# U-Net as a deep learning-based method for platelets segmentation in microscopic images

**DOI:** 10.1101/2024.08.23.24312502

**Authors:** Ajay Kumar, Charlie A. Coupland, Tania F. Vaz, Will Jones, Rubén Valcarce-Diñeiro, Simon D. J. Calaminus, Eva Sousa

## Abstract

Manual counting of platelets, in microscopy images, is greatly time-consuming. Our goal was to automatically segment and count platelets images using a deep learning approach, applying U-Net and Fully Convolutional Network (FCN) modelling. Data preprocessing was done by creating binary masks and utilizing supervised learning with ground-truth labels. Data augmentation was implemented, for improved model robustness and detection. The number of detected regions was then retrieved as a count. The study investigated the U-Net models performance with different datasets, indicating notable improvements in segmentation metrics as the dataset size increased, while FCN performance was only evaluated with the smaller dataset and abandoned due to poor results. U-Net surpassed FCN in both detection and counting measures in the smaller dataset Dice 0.90, accuracy of 0.96 (U-Net) vs Dice 0.60 and 0.81 (FCN). When tested in a bigger dataset U-Net produced even better values (Dice 0.99, accuracy of 0.98). The U-Net model proves to be particularly effective as the dataset size increases, showcasing its versatility and accuracy in handling varying cell sizes and appearances. These data show potential areas for further improvement and the promising application of deep learning in automating cell segmentation for diverse life science research applications.

**Author Summary:** Deep Learning can be used with good results for automatic cells images segmentations, reducing the time applied by scientists to this task. In our research platelets images were automatically segmented and counted using by applying U-Net and Fully Convolutional Network (FCN) modelling. Data preprocessing was done by creating binary masks and utilizing supervised learning with ground-truth labels, after data augmentation. U-Net surpassed FCN in both detection and counting measures in a smaller dataset. The U-Net model proves to be particularly effective as the dataset size increases, showcasing its versatility and accuracy in handling varying cell sizes and appearances. Our study shows potential areas for further improvement and the promising application of deep learning in automating cell segmentation for diverse life science research applications.

## Introduction

In recent years, cell segmentation has emerged as a critical component in numerous research fields, including bioinformatics, cell biology, and computational biology [1–3]. Deep convolutional neural networks (CNNs) have revolutionized visual recognition tasks, outperforming traditional methods across various domains [4]. By utilizing CNNs, deep learning algorithms have demonstrated the ability to accurately identify and count cells in biomedical images [5]. However, the conventional use of CNNs in classification tasks does not fully address the complexities of cell segmentation in microscopy images, where pixel-level localization is crucial [6].

Cell segmentation, the process of delineating cell boundaries in microscopy images, is a critical step for morphological analysis and downstream quantification of biological structures. Since 2015, a range of deep CNN architectures have achieved breakthrough results on standard cell segmentation benchmarks [7]. Early networks like U-Net by Ronneberger et al. [8] introduced a symmetric encoder-decoder structure to propagate multi-scale contextual information, which became highly influential. Other top designs utilized pre-trained classification backbones like Visual Geometry Group by Simonyan and Zisserman [9] or Residual Networks by He et al. [10], to effectively initialize deep models. More recent techniques further incorporated elements like atrous convolutions by Chen et al. [11] and generative adversarial training [12] to capture both local details and global consistency. Powered by ever larger annotated datasets, these latest CNNs have surpassed human experts on nuclei segmentation and approaching expert inter-observer agreement on challenging cell contouring tasks [13,14].

However, substantial obstacles prevent the real-world adoption of deep learning segmentation tools. Cell images exhibit high appearance variability under different experimental conditions, with artifacts like missing cellular boundaries, that easily confuse the models [15]. Complexities such as cell-cell interactions, for example overlapping cells or cell-background interactions, also pose difficulties [16]. Such data heterogeneity issues combined with label noise and inconsistencies during manual segmentation, poses several segmentation challenges [17,18]. Furthermore, while state-of-the-art results are reported on some curated test images, deep networks frequently fail to generalize across different imaging setups without extensive retraining [19].

To overcome these limitations, recent works have proposed techniques to improve model robustness. Generative and reconstructive approaches to incorporate unlabelled data during training can enhance generalizability [20]. Assessment of remaining errors to guide annotation and data augmentation can mitigate dataset bias [21]. Through focused incorporation of these sophisticated regularization, adaptation, and interaction techniques, deep CNNs may eventually fulfil their promise for practical automated cell segmentation [22,23].

Nowadays, both U-Net, as a particular type of FCN and FCN in general are known as CNN architectures to be employed in microscopy and biomedical image analysis, with U-Net being a particular type of FCN [24,25]. While FCN utilizes a classification network like ImageNet by Krizhevsky et al. [26], and U-Net was built on fully convolutional network (FCN) with hourglass topology [8,24]. Semantic segmentation framework is based on a bounding box-based segmentation pipeline that extracts the foreground from a given region of interest. It focused on image local patterns and extracted complex image information at various scales. It has proven to be successful in biomedical applications and has gained popularity in many research studies in cell detection [5,27] and cell segmentation [28,29].

The recent growing of deep learning applications for microscopic analysis is revolutionizing the process of classifying, counting and segmenting cells [30]. These tasks which traditionally were performed by humans and are very time consuming, have a high potential of success to be fully automated thought deep learning algorithms with good results [31]. Furthermore, manual segmentation also introduces a high degree of user subjectivity and variability which may have an impact on the experimental results obtained [31]. Therefore, this research aims to build an automated system for platelets segmentation and respective size determination, on microscopy images, by creating a mask that allows the platelets detection and counting.

## Results

### Model evaluation

Experiments with the smaller dataset (293 images) where performed both with FCN and with U-Net, while only the two bigger datasets (1172 and 4688 images) were used with U-Net. Experiments with FCN were abandoned after experiments with the smallest data set due to its inferior comparative performance. Different methodologies of threshold were used for the two types of networks. FCN used a method of segmentation by the Sobel operator, while U-Net used binarization of the image to create ground-truth masks for segmentation.

### FCN model evaluation

Data pre-processing within the FCN model generated masks as segmented images from Sobel operator at a kernel or threshold value of 7, as shown in Fig 1. As it can be seen the Sobel operator tends to enhance the edges of the platelets [32].

**Fig 1.**
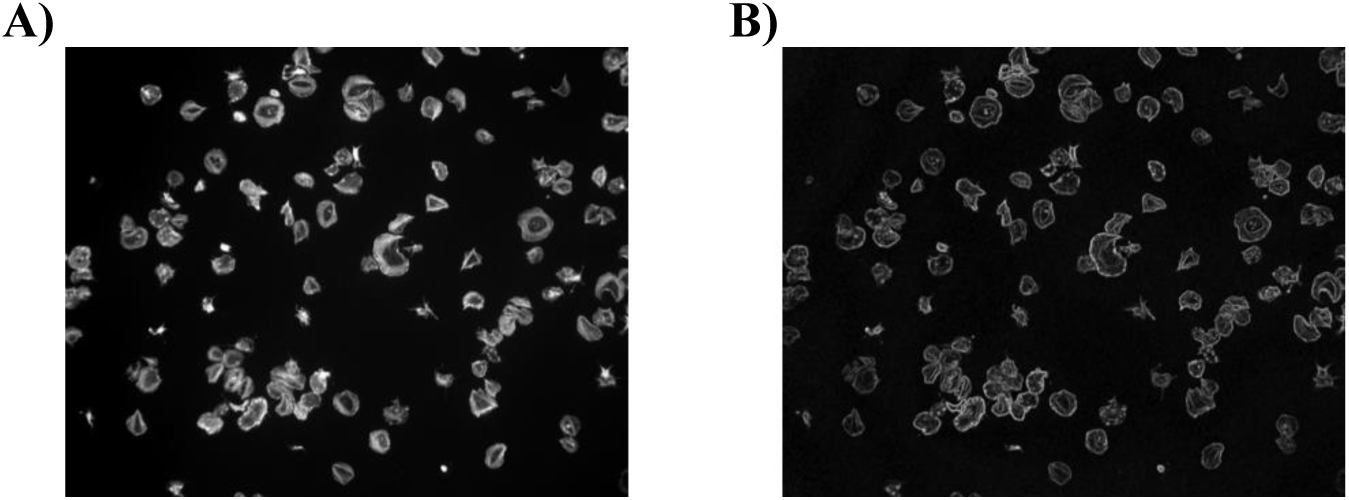
Identification of Sobel Segmented Images within FCN model. Platelets (2×10^7^/ml) were spread on fibrinogen for up to 45 minutes, before fixation, staining, and imaging using a Zeiss Axiovert Fluorescent microscope (oil x63 NA 1.4 objective). (A) Image is representative of control conditions. (B) Representative image segmented by enhanced Sobel operator.

The FCN model was evaluated with the complete iteration of 10 epochs utilizing a processing time of 1206 seconds and resulted in an accuracy of 0.81, reaching an Area Under the Curve (AUC) of the Receiver Operating Characteristic (ROC) of 0.71. This and the loss function can be seen in Fig 2. For visually inspecting the cell segmentation and the model’s performance, the ground truth masks were compared with the masks predict-ed by the model. But the model failed to demonstrate or produce predicted cell counts as it resulted in MPE of 55.44%.

**Fig 2.**
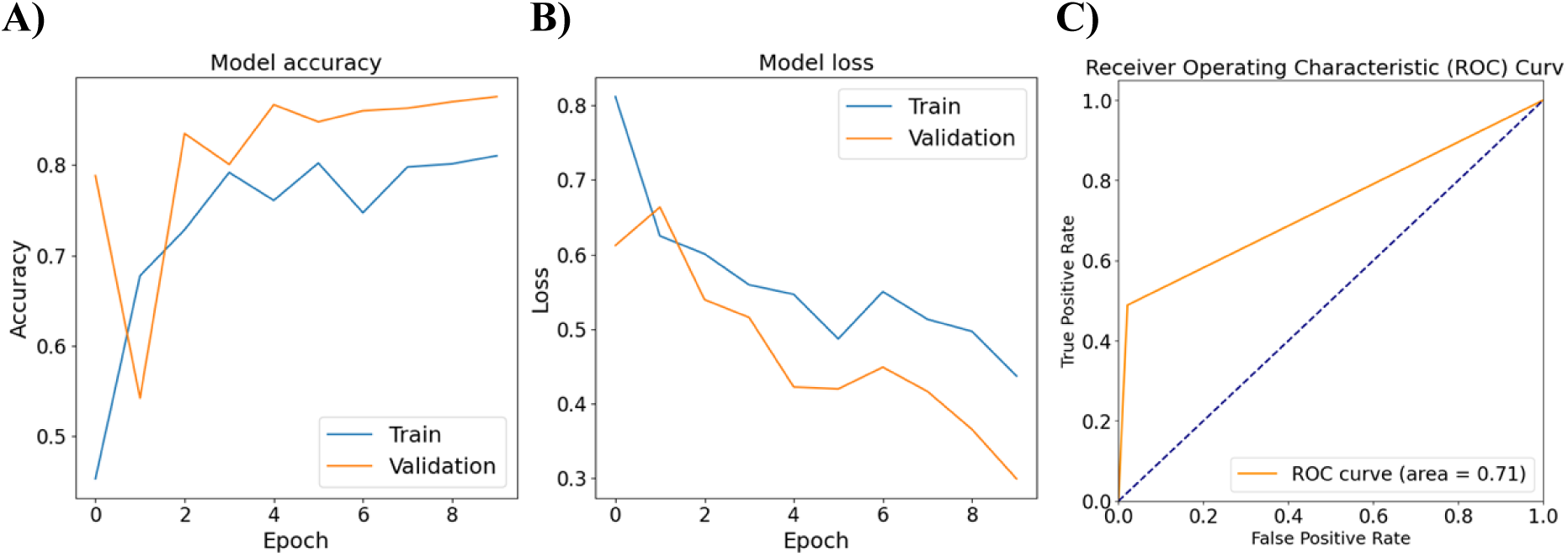
Plots of FCN model evaluated on 293 images. (A) training and validation loss; (B) accuracy; and (C) AUC-ROC.

### U-Net model evaluation

In the U-Net model instead of the Sobel operator, binary masks of the platelets images, named ground-truth masks were created preprocessing using a threshold of 25 for image binarization. An example of the ground truth mask, and of the image from which were created are shown in Fig 3.

**Fig 3.**
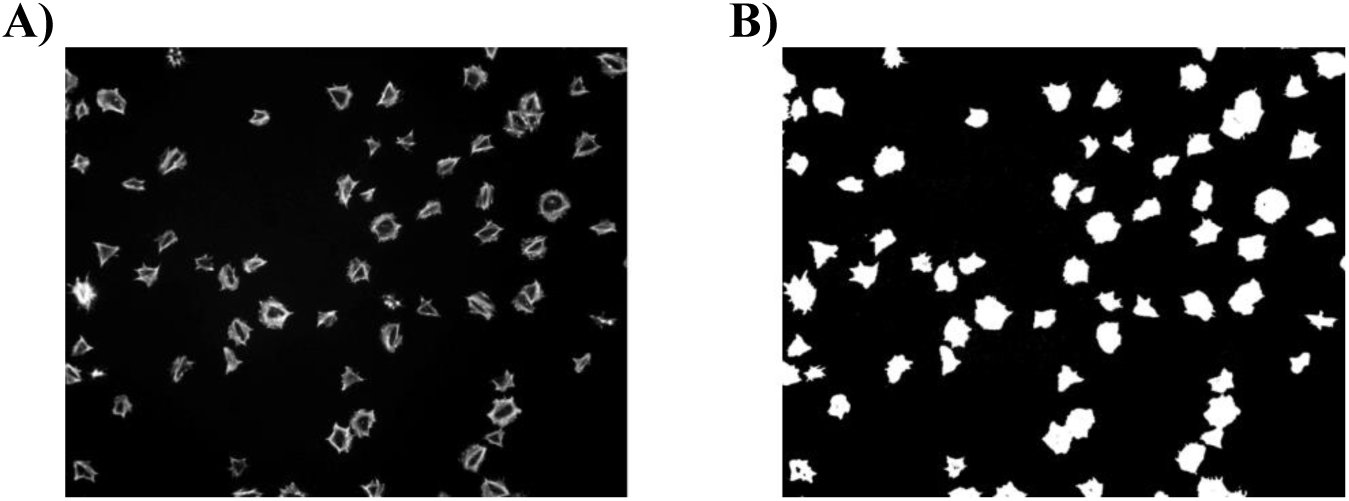
Identification of ground truth marks within U-Net model. Platelets (2×107/ml) were spread on fibrinogen for upto 45 minutes, before fixation, staining and imaging using a Zeiss Axiovert Fluorescent microscope (oil x63 NA 1.4 objective). (A) Image is representative of control conditions. (B) Representative image with the corresponding ground-truth masks.

The U-net model was pre-trained with generated masks and a complete iteration of 10 epochs was monitored on both the training and validation datasets resulting in an accuracy of 0.96. A lower MAE of 2.6% suggests that model’s predictions are close to the true values, reflecting accuracy in pixel-wise predictions, as shown in Fig 4.

**Fig 4.**
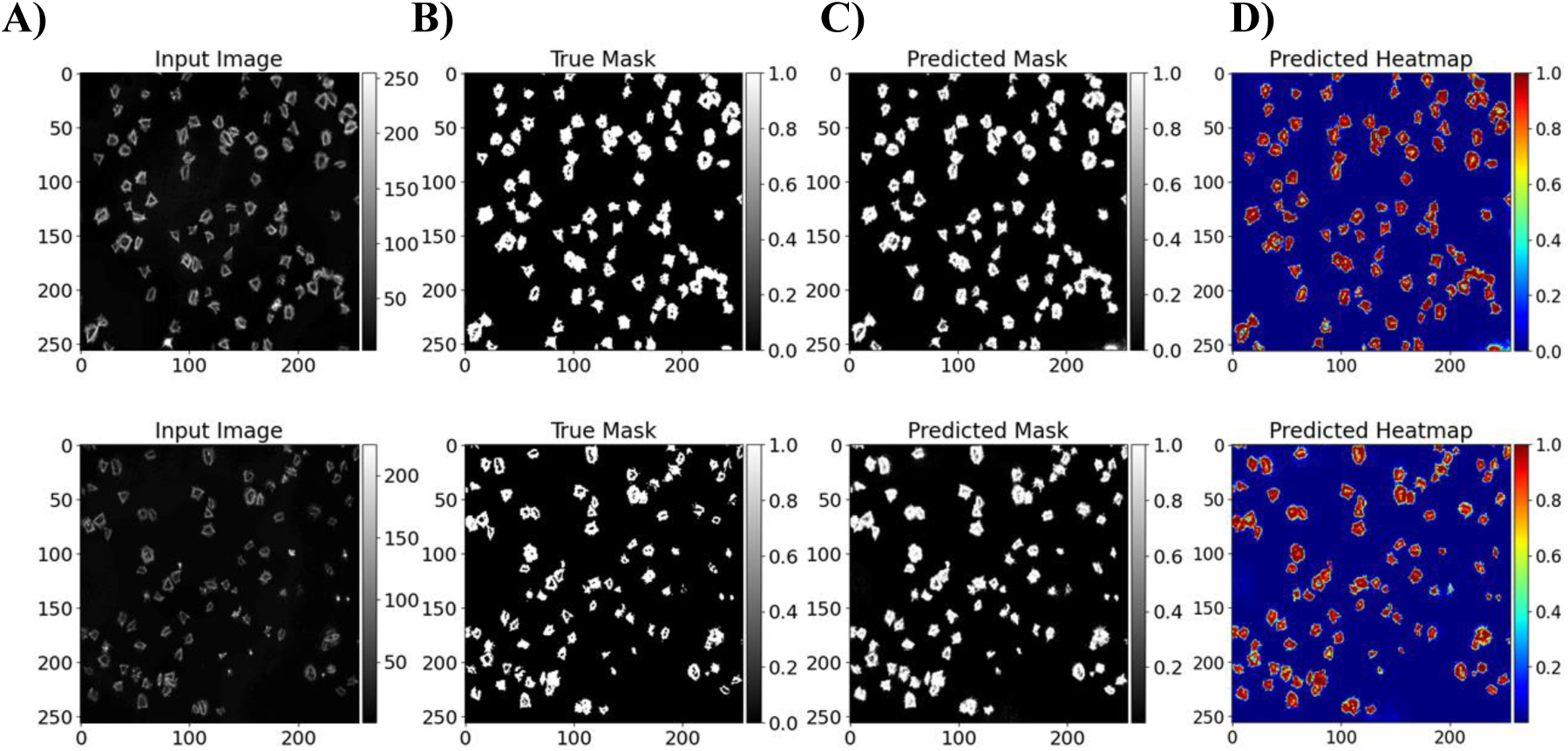
Identification of platelet segmentation with U-Net model. Platelets (2×107/ml) were spread on fibrinogen for up to 45 minutes, before fixation, staining and imaging using a Zeiss Axiovert Fluorescent microscope (oil x63 NA 1.4 objective). (A) Image is representative of control conditions; (B) representative image with ground-truth masks; (C) predicted images from U-Net model; (D) corresponding heat maps. All images have a dimension of 256×256 pixels.

Lastly, the U-Net model was evaluated by rotating 1172 images into specified degrees of 90, 180, and 270, creating a combined dataset of 4688, and their corresponding masks were generated at threshold of 25. The training accuracy reached 0.98 and the vali-dation loss continued to improve across epochs reaching an AUC of the ROC of 0.99, as shown in Fig 5.

**Fig 5.**
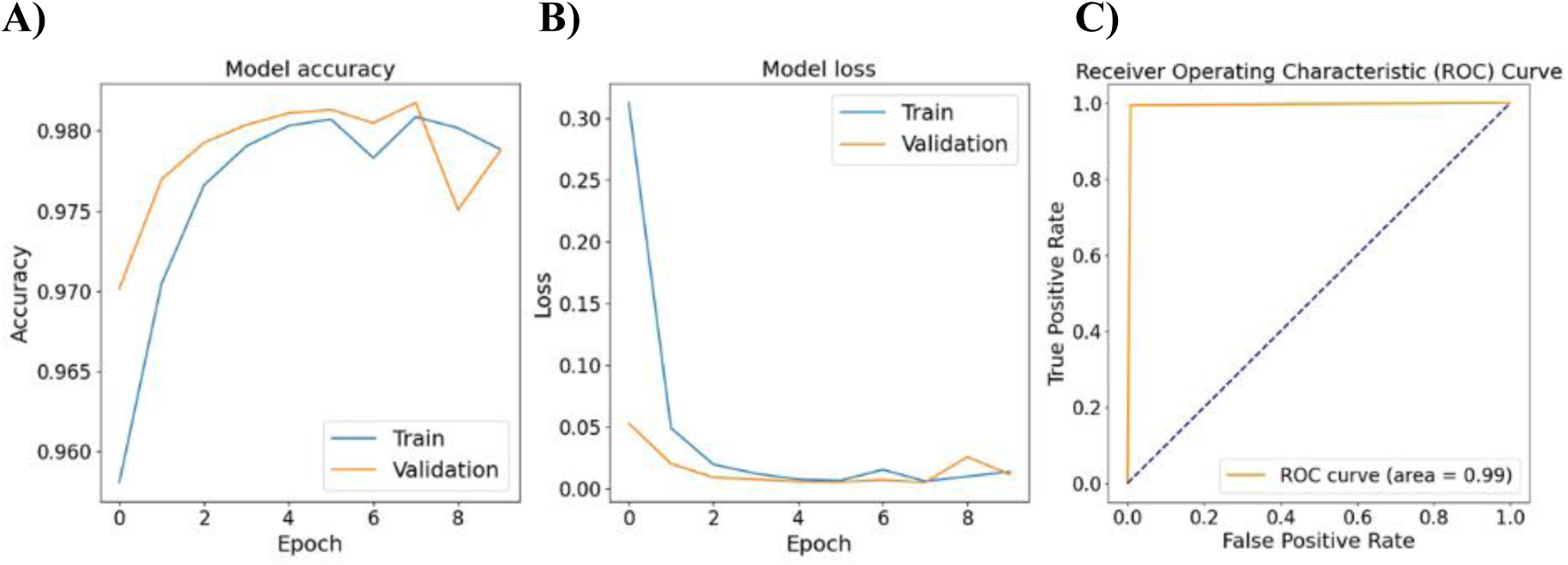
Plots of the U-Net model evaluated on 4688 images. (A) training and validation accuracy, (B) loss and (C) AUC-ROC.

### Cell counting by U-Net

Following the training of the U-Net, the subsequent phase involved cell count drawn from a dataset encompassing 4688 images. The training employed a L2 loss function, incorporating aleatoric uncertainty for cell counts (Fig 6), and optimization was carried out using the Adam optimizer with a learning rate of 1*E*-4 and a batch size of 8 (Fig 8). The estimation of cell sizes in the predicted masks, as shown in Fig 7, for each segmented cell (region) was calculated in terms of number of pixels it occupies at region (bounding box size) with a threshold of 50.

**Fig 6.**
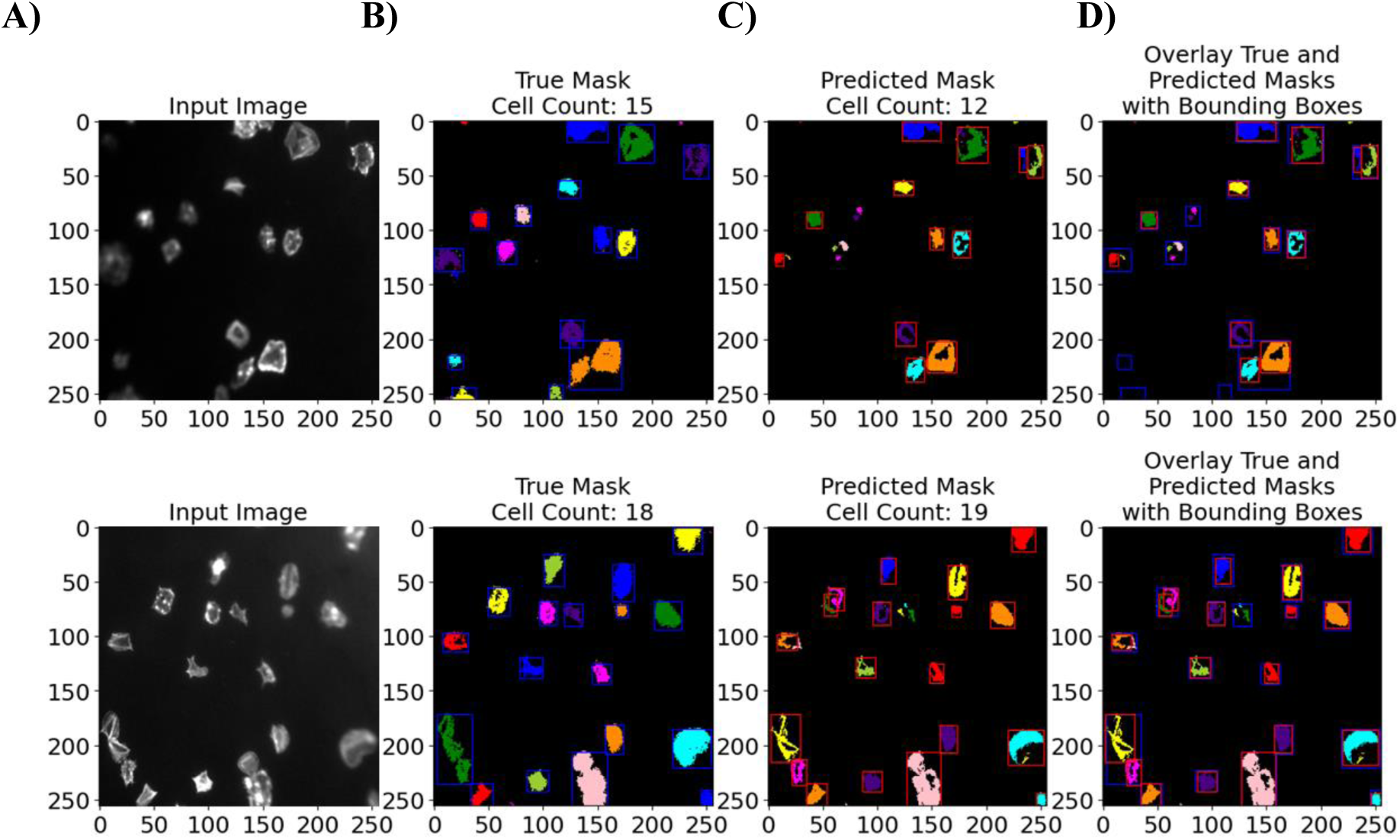
Two examples of Sample of segmentation results of images from U-Net model cell counts of true and predicted masks. (A) Original input images, (B) ground truth masks, (C) predicted masks, and (D) corresponding overlay. All images have a dimension of 256×256 pixels.

**Fig 7.**
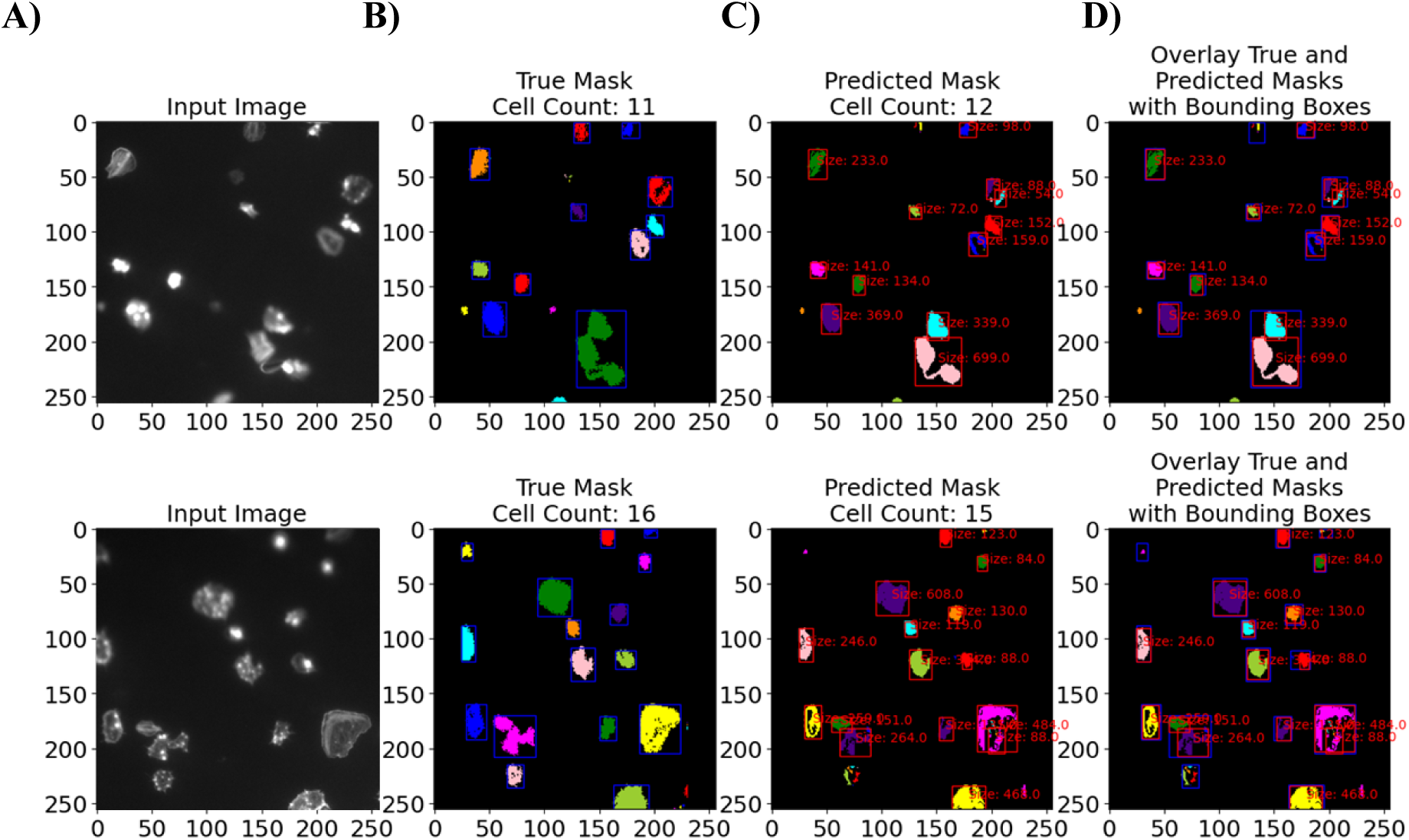
Two examples of Sample of segmentation results of images from U-Net model with cell size in number of pixels. (A) Original input images, (B) ground truth masks, (C) predicted masks, and (D) corresponding overlay. All images have a dimension of 256×256 pixels.

**Fig 8.**
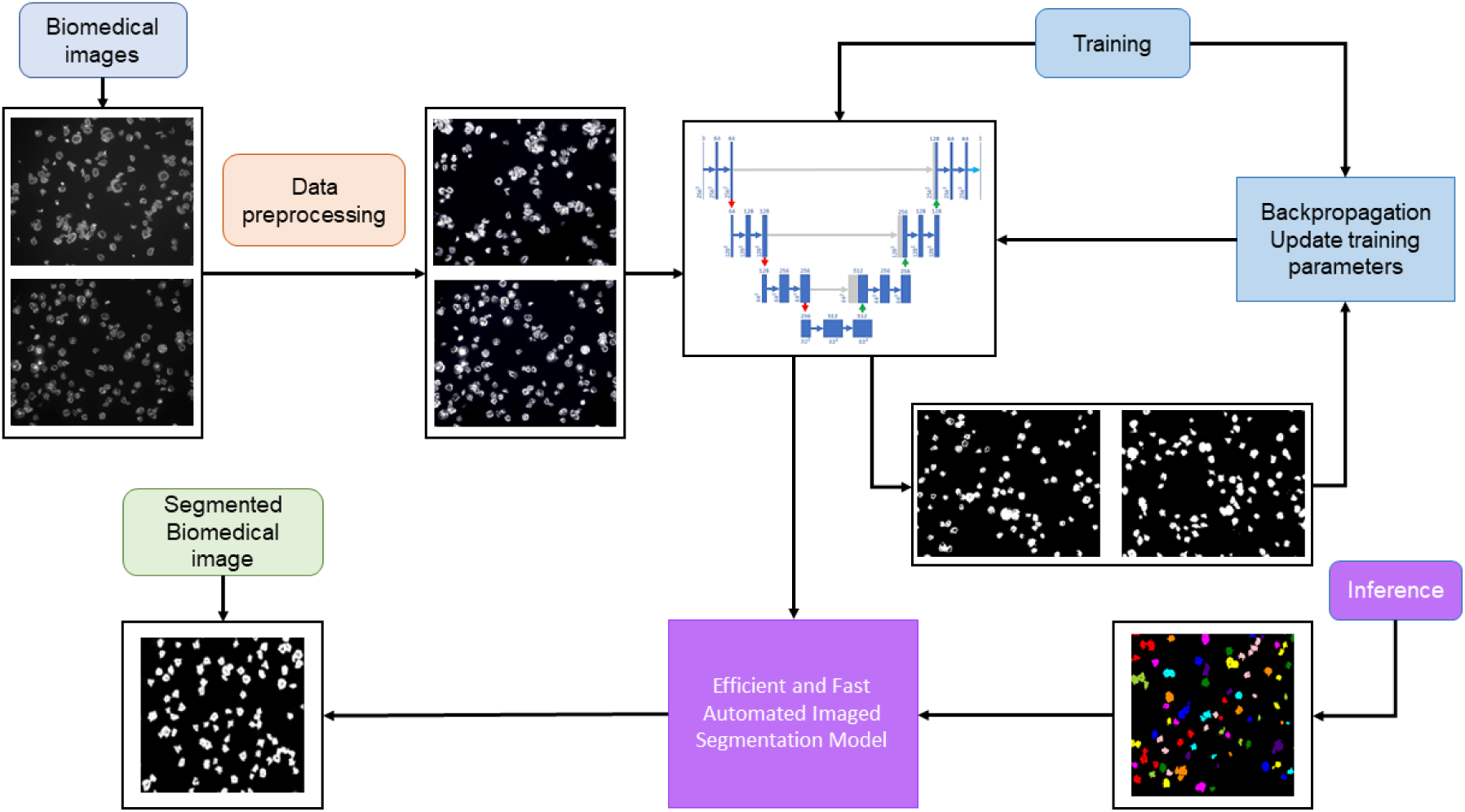
Diagram of the methodology approach for segmenting platelets in biomedical images using the U-Net. Adapted from [54].

### Cell segmentation and quantitative evaluation by U-Net

The outcomes derived from the evaluation of U-Net models on the three dataset groups indicated a very positive correlation between the size of the dataset and the positive results, achieving higher values for the bigger dataset, as it can be seen in Table 1. In the biggest dataset the maximum values are accuracy of 0.98, recall 0.98, precision of 0.99, IoU of 0.99 and Dice of 0.99. Additionally, a concurrent reduction in both dice loss and sparse categorical cross-entropy loss was observed, as the employed loss functions exhibited a robust interdependence and displayed an inverse relationship with accuracy (Figs 5A and 5B). Following the specified criteria, the most favourable U-Net configuration [33] involved five encoding and decoding blocks. This assessment also extended to the examination of the number of filters in the initial encoding block, revealing a doubling of filters with each subsequent block and a corresponding halving with each decoding block. The optimum number of filters, within the investigated parameters, was identified as 64 in the first encoding block. This U-Net has been used for other types of segmentation tasks [33] but was revealed more successful in the application to platelets segmentation than another U-Net model initially tested, and which is more commonly used in cell segmentation [8].

**Table 1.**
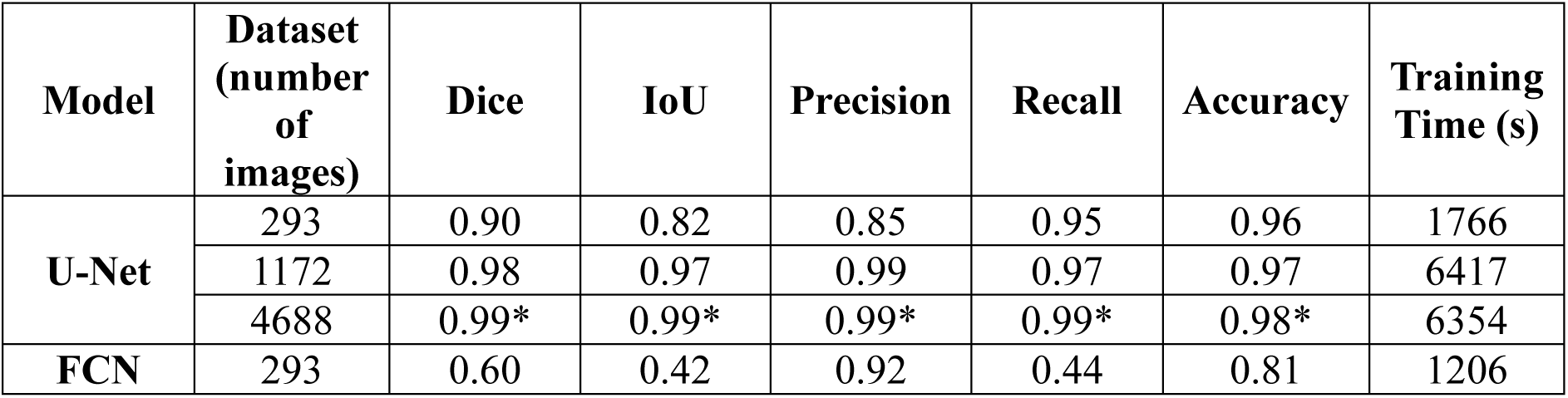
Cell segmentation performance results computed for each model.

Table 1 elucidates the performance outcomes, throughout several metrics, of cell segmentation derived from the evaluation of the test set for each model. The evaluation was conducted across distinct datasets characterized by varying numbers of images, with the U-Net model serving as the segmentation architecture.

Across the dataset comprising 293 images, the U-Net model achieved a Dice coefficient of 0.90, indicating a substantial agreement between the predicted and ground-truth segmentation masks. The IoU, measuring the overlap between the predicted and true segmentations, was 0.82. The precision, reflecting the positive predictive value, was observed to be 0.85, while recall, gauging the model’s ability to capture all positive instances, exhibited a value of 0.95. The overall accuracy, encompassing both true positive and true negative predictions, was 0.96. It is worth noting that using the optimum threshold of 0.25 assures huge cut-offs and enforces only detections with high confidence, as this was the only value in which all the platelets were correctly included in the binarization of the im-aging. A too low threshold would increase the areas beyond the platelets area, while a too high one would confuse darker platelets as background. Although desirable, this behaviour increases false negatives, as fewer platelets are spotted resulting in accuracy down-fall, with the impact of false negatives being twice as large as it is in Dice of 0.90, explaining the disparity between these two metrics. The single significant exception is accuracy, which U-Net architectures excels at. This is most likely owing to an “over-detection” tendency. Nonetheless, the FCN counterparts outperform this tendency by significantly im-proving accuracy and precision reported at 0.81 and 0.92, respectively.

As the dataset size increases to 1172 and 4688 images, the U-Net model demonstrated notable improvements in performance metrics. The Dice coefficient increased to 0.98 and 0.99 respectively, indicating enhanced segmentation agreement, while IoU rises to 0.97 and 0.99, depicting increased overlap between predicted and true segmentation. The consistently high values across various evaluation metrics sustained the U-Net model’s effectiveness in handling datasets with varying cell sizes and appearances, as reported in Table 1. The model exhibited further refinement in segmentation due to a series of deconvolutional layers that reconstructed the output image from the extracted features as the training data increased. In contrast, FCN models do not have analogous shortcut paths to retain and fuse low-level information through the network architecture. IoU of 0.42 proves that as sequential encoder-decoder flows, FCN faces more challenges restoring spatial de-tails from compressed latent bottles when built on smaller datasets, as it was seen from the results of the comparison of both models in the 293 images set where both were tested. For this smaller dataset, U-Net model’s MAP of 0.99 denoted exceptional precision across the dataset, suggesting a minimal number of false positives and high relevance in the predicted outcomes. MAE of 0.002 and MPE of −0.050 indicated a close alignment between the model’s prediction with a negative sign showing a slight underestimation on average. For the FCN Model, the results suggest that it performs reasonably well in terms of precision, with MAP scores around 0.8365. However, there is room for improvement in reducing the absolute and percentage errors in pixel-wise predictions, as indicated by the MAE and MPE values of 0.1828 and 0.5544 respectively.

## Discussion

Deep Learning use for imaging classification, segmentation and counting has some advantages over this work being done by humans. First convolutional neural networks are more consistent than humans, as they will (1) classify images identically each time, (2) do not introduce differences in the procedure (3), are a great time saver [33]. Given all the improvement possibilities for imaging classification, segmentation and counting, it is of crucial importance to find suitable methods to support or replace humans in these tasks where it is possible [34,35].

From our research we are led to believe that the U-Net model could be very promising to aide in effective analysis of platelets microscopic images. This U-Net [33] model uses the notion of deconvolution by [4,33] analysis and synthesis. The analytical path follows CNNs structure as shown in Fig 8 and the expansion step of the synthesis path consisted of an up-sampling layer followed by a deconvolution layer. It is found that the most essential aspect of U-Net is the ability to create shortcut connections between layers of equal resolution in the analysis path and the expansion path. These connections supply the deconvolution layers with critical high-resolution features [36,37].

The studies undertaken in this research stand on the implementation of two specific design choices which were found to significantly enhance the performance of the model. Firstly, the incorporation of ground truth masks, and second the application of a U-Net model. The incorporation of ground truth masks penalizes errors occurring on cell boundaries and in densely populated regions, proving to be instrumental in promoting precise segmentation, particularly in scenarios involving closely situated objects [38,39]. Similarly to what was reported in the bibliography [40,41] in our comparative analysis the U-Net model stands out as the most effective network outperforming the FCN (Table 1) across all performance metrics apart from the precision and training time, when both U-Net and FCN were applied to the smallest dataset. Given this difference in performance, only U-Net was applied to the bigger datasets with excellent results, in all metrics, and without much computational time added. It is important to note that as the dataset in-creased four-fold in complexity the processing times of the U-Net model remained similar (at 6417 and 6354 sec respectively). This a very advantageous characteristic when searching for a model to train [42].

This success seems to be due to the combination of (1) ground truth masks and (2) U-Net architecture which demonstrated high accuracy in cell count predictions and adheres to the conservative counting requirement that underscores that precise cell counts are a result of accurate object detection rather than a mere balancing effect between false positives and false negatives [43].

In our work instead of applying a standard U-Net five-layer convolutional module already in use for cell segmentation [8], a four-layer module was used to meet the segmentation task and avoid excessive parameters [33].

Here an encoder with a succession of convolution and max pooling layers characterized the network, with a mirrored sequence of transposed convolutions in the decoding layer. The U-Net model learns the crucial features of the images after encoding, and to segment the image needs to decode them. Each convolutional block in the decoder has the same settings as those in the encoder. After each convolutional block, the image is up-sampled twice using bilinear interpolation to make it larger. Then, a skip connection links it to the corresponding feature map in the encoder. It utilizes a 1×1 convolutional layer after last set of decoder blocks to construct the final segmented image and for the conversion of RGB to grayscale.

The layer of convolution network in the FCN model is a three-dimensional data array, with each layer representing an image with height x width x depth pixels and colour channels [24]. The image is the initial layer, with receptive fields representing the image’s positions. Convolution, pooling, and activation functions operate on local input regions and are based on translation invariance. The inclusion of bounding boxes around regions facilitated the quantitative assessment of segmentation accuracy (Fig 6).

Additionally, when considering uncertainty predictions, over 80% of ground-truth counts were found to fall within the model’s predicted 95% confidence interval across our 750-image test (examples showed in Fig 6). This visualization is invaluable for understanding the segmentation performance, assessing the accuracy of cell delineation, and providing insights into potential areas for improvement. The inclusion of cell sizes and not only of the cells counting, enhances the interpretability of the segmentation results by providing quantitative information about the segmented platelets within the predicted masks (Fig 7).

The absence of foreground masks for out-of-focus images in the dataset hinders counting performance, suggesting the potential for enhancement through the inclusion of such masks. However, challenges may persist [16,44,45], particularly in cases of overlapping cells (platelets) [46,47], a difficulty even confounding human experts. To address this, a plausible strategy involves incorporating the original image as supplementary input to the counting network. Additionally, another approach could entail utilizing randomly cropped image patches and robustly estimating counts by averaging density across multiple patches, akin to the methodology proposed by Oñoro-Rubio and López-Sastre [48].

Notably, similarly to other works the strategic enhancements we introduced in comparison to the original U-Net architecture, specifically the integration of a learned transformation and the inclusion of a residual block with 3 × 3 filters, seem to significantly contribute to the model’s superior performance [49,50]. Lastly, it becomes evident that even instances of misidentification possess a certain degree of subjectivity, residing within the nuanced boundaries of interpretability for borderline cases (examples showed in Fig 6).

Finally, it was shown how aleatoric losses can be used to estimate uncertainty in cell counting for failure cases where ground-truth is outside of some acceptable tolerance [51]. Our work is limited by the requirement of annotated datasets, in which the bias of the labelling can be introduced.

Similarly to the U-Net model, FCN’s architecture consists of multiple convolutional layers to collect features from input data, and pooling layers minimize the spatial dimensions of the data to capture the most significant information. But given our results when comparing it with the U-Net model it was shown not to be the most optimum model.

In summary, the proposed approach, with the U-Net and ground truth masks, has demonstrated its robustness to be applied for automating prevalent operations across various life science research applications. Consequently, this strategy holds the potential to yield significant advantages in terms of expediting studies and mitigating operator bias, both within individual experiments and across diverse experimental contexts.

## Material and Methods

This study proposed the implementation of the U-Net and FCN models for accurately segment platelets, in microscopy images. Platelets are dense and adherent cells, causing extra difficulties in the segmentation task [52,53]. In Fig 8 is depicted the overall procedure devised using U-Net for segmenting, counting, and calculating the area of the platelets, as this was the best performing system. All the procedures begun with the pre-processing of original microscope cell images, and preparation of the databases by augmentation of the original datasets. The preprocessing of the images was done primarily by adjusting image size of the images, followed by segmentation procedures. After detecting the platelets, the final counting is obtained as the number of connected pixels in the post-processed output. The study design decisions, such as the chosen threshold, were all aimed at reducing false negatives and promoting accurate segmentation, and images quality highly influences the training of the network, and the segmentation results possible to be achieved by it.

### Dataset

The suggested framework was assessed using a dataset gathered by the Centre for Biomedicine, Hull York Medical School, University of Hull, UK [55]. The original database includes 293 microscopy images that have been carefully classified by skilled experts. These datasets showed platelets clustered together with low-contrast cell borders. Cell size and appearance varied between datasets. The first dataset consisted in 293 microscopy cell images of human blood platelets after different treatments: with Zinc, Milrinone, and Mil-rinone + Zinc (in a total of 299, excluding five duplicates and one blank image). From the ethical perspective, no image annotation tool was used as the dataset does not contain an-notations or labelling. To increase the dataset size, were created two datasets with 1172 and 4688 cell images by data augmentation (splitting and rotating the original images). The original dataset of 293 images was an 8-bit 3-channel jpeg of 2752×2208 pixels each (Fig 9). The dataset of 1172 images were created by splitting 293 images into 4 quadrants resulting in 3-channel 1376 x 1104 pixels each. Applying rotating methods, the dataset was increased from 1172 to 4688.

**Fig 9.**
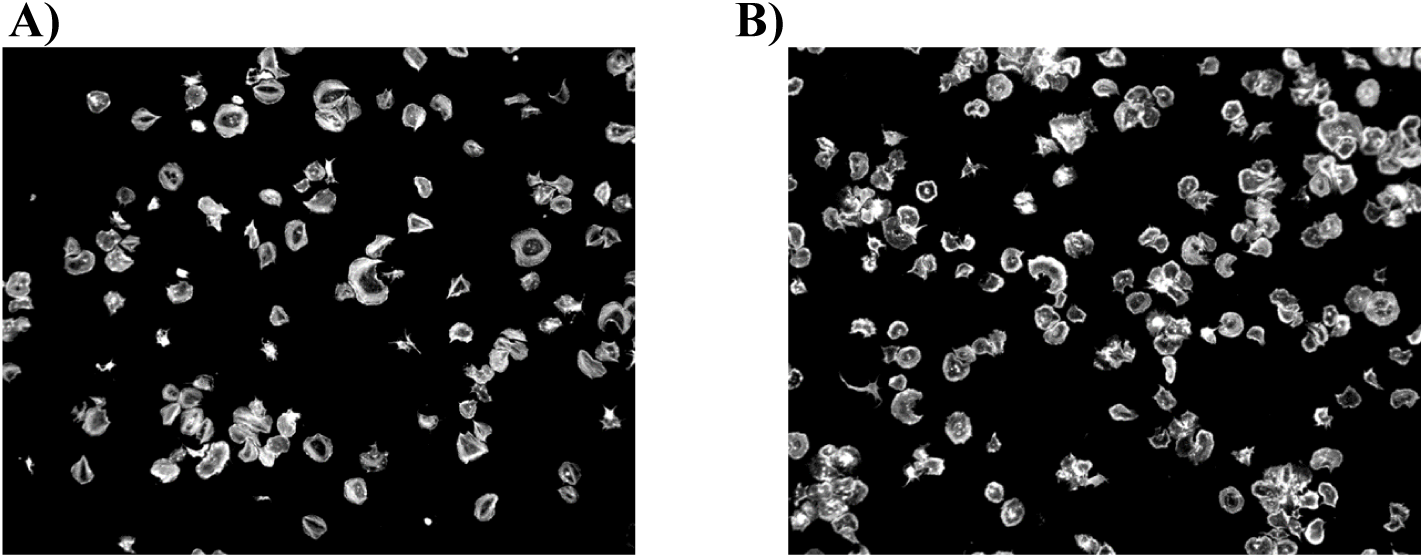
A and B). Two different samples of microscopy cell images from the original dataset. Images with 2752×2208 pixels.

### Data preprocessing

Firstly, the dataset of 293 images was inspected and duplicates were removed to create this new cleaned database. Following, contrast was enhanced using Contrast Limited Adaptive Histogram Equalization (CLAHE) with clip limit of 3, which is a contrast enhancement technique that prevents over-amplification of noise. The data augmentation technique introduced new patterns into the training dataset, which made the training procedure more resistant to over-fitting, and was applied with randomized rigid geometric changes, scaling, and colour values (grey), where each training sample was rescaled, and then randomly spun before flipping it. A standard split of 80-20 train/test was used for all the final models, with the different tested datasets.

For both FCN and U-Net models masks were created for the segmentation but the procedure to create these masks was different. For the FCN model, masks were created as segmented images from Sobel operator at kernel value of 3, 5, 7 and 9 as appropriate. A larger kernel size increases sensitivity to broader edges but might reduce localization accuracy for finer details. The function in the sobel operator calculates the gradient magnitude by taking the square root of the sum of squared horizontal and vertical Sobel responses (sobel_x and sobel_y respectively) providing a combined measure of edge strength in both directions.

For visually inspecting the cell segmentation performed by the FCN Model and the model’s performance, the ground truth masks were compared with the masks predicted by the model. For cell counting from the segmented images, multiple threshold values of 0.10, 0.15, 0.25, and 0.5 were tested for minimum area or region of interest for creating bounding boxes. For the U-Net model ground-truth masks were created by binarization of the images.

This binarization happened from the threshold value which allowed all the cells to be binarized. As well here several binarization threshold values were tested, namely 0.05, 0.15, 0.25, 0.30 and 0.50. 0.25 was considered the optimum threshold value for creating the ground truth masks as all the platelets would be binarized in a close area.

In the training phase, a supervised learning framework used the ground-truth la-belled images, as samples of desirable outputs that the model should learn to generate. In the case of image segmentation, such targets take the form of binary images (masks), with white (0) and black (1) pixels, representing the objects to segment and the background, respectively (Figure 1). To the cleaned images was then applied a second threshold using automatic histogram shape-based algorithms. Region properties were calculated, and bounding boxes were drawn around regions with an area exceeding the specified second threshold applied of 0.5 to match the true mask and to eliminate the smaller particles or noise.

### Model architecture and training

FCN architecture consists of an encoder-decoder structure. The encoder extracts features from the input images through convolutional and pooling layers, while the decoder up-samples these features to generate pixel-wise predictions. Skip connections were incorporated to preserve spatial information during upsampling.

The FCN architecture chosen comprised 2 layers of 4 convolutional blocks with 64 and 128 filters in both the encoder and decoder section, with 2 max-pooling layers in the encoder, 2 Up-sampling layers and 2 concatenation layers (one for each decoding block), and one final convolutional layer with a sigmoid activation function. The model was compiled using the Adam optimizer and binary cross-entropy loss function and the model was trained on the smaller dataset. A validation split of 20% was used to monitor the model’s performance during training.

The model was evaluated on training and validation datasets with the complete iteration of 10 epochs but abandoned due to less successful results than the U-Net model ap-plied. The U-Net model used in our study [33], started with a defined input layer, accommodating image size of 256×256 pixels with 3-color channel. For deeper feature extraction in encoder portion, a series of convolutional layers with 64, 128, and 256 filters of size 3×3 interspersed with rectified linear unit (ReLU) activations and max-pooling operations progressively reducing spatial dimensions, followed by a middle bottleneck layer of 512 filters to capture contextual information and a decoder segment with similar layers as encoder segment, which progressively up samples feature maps and concatenates them with feature maps from the corresponding encoder layers, enhancing localization precision.

Each of these convolutional blocks in the encoder, employed edge filling for each convolutional layer to maintain the feature map and the ReLU function, and is expressed in equation 1.

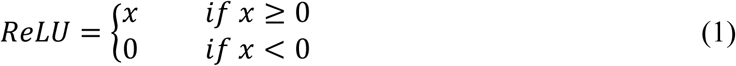

The final layer employed a sigmoid activation, followed by Adaptive Moment Estimation (Adam) optimizer with learning rate of 1*E*-4, and binary cross-entropy loss which quantifies the dissimilarity between predicted and ground-truth segmentation maps.

In this scenario, image segmentation required unannotated data with ground-truth labels resulting in an unsupervised or weakly supervised image segmentation approach, and the construction of a loss function capable of assessing the quality of segments or clusters of pixels is the key difficulty. The model was compiled with binary cross-entropy as the loss function. In our study the U-Net model (Fig 10) was trained and tested on 234 and 59 cell images and ground-truth masks, respectively.

**Fig 10.**
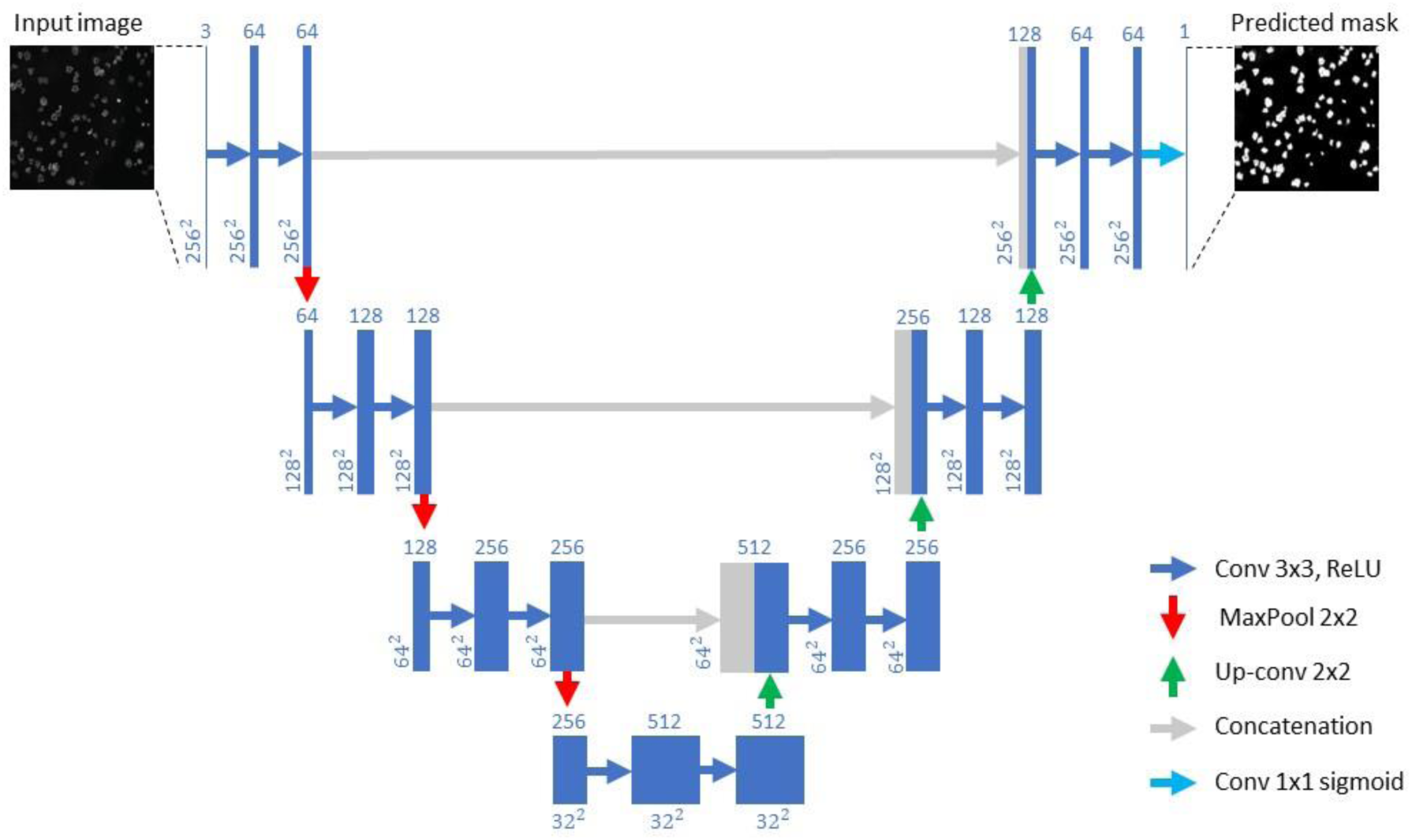
U-Net applied architecture. Adapted from [33].

### Metrics for model performance evaluation

Intersection over Union (IoU) or the Jaccard Index (J), is a widely used metric in semantic segmentation, where A and B represent the true and predicted segmentation maps, respectively (Equation 1), and Dice (Equation 2).

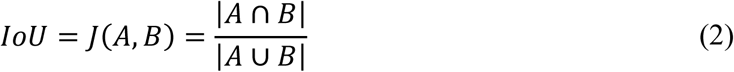

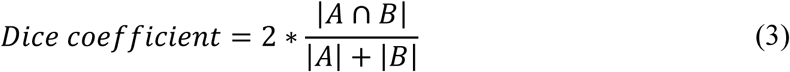

To calculate the overall detection of platelets (True Positive (TP)), it is assumed that the system properly detected more than 50% of the pixels. Precision (Equation 4), recall (Equation 5) and accuracy (Equation 6), were used for reporting the accuracy of image segmentation techniques. For pixel-wise comparison between the expected and the achieved were used the Mean Absolute Error (MAE) (Equation 7), Mean Percentage Error (MPE) (Equation 8) and Mean Average Precision (MAP) (Equation 9) were calculated.

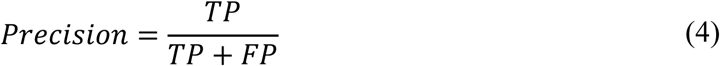

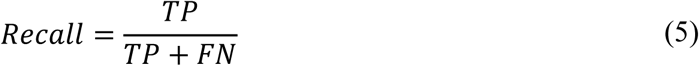

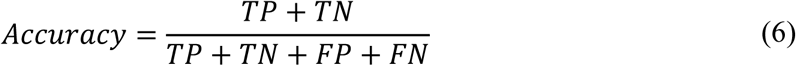

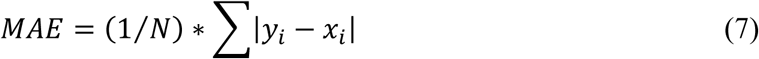

Where *N* is the number of samples, and |*y*_*i*_ − *x*_*i*_| the error in absolute values.

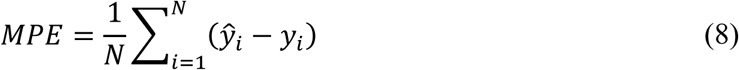

Where *N* is the number of samples, ŷ_*i*_ is the forecasting value, and *y*_*i*_ is actual load value.

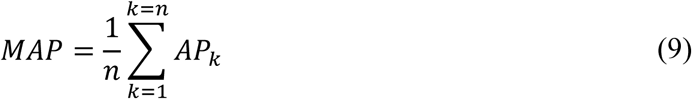

Where *n* is equal to the number of classes and *AP*_*k*_ the average precision of the class *k*.

### Software and hardware

The experiments were conducted on a system running Windows 11 Home 23H2. Data preprocessing was performed using Python 3.11.3 with scikit-learn (1.2.2) library. The deep learning models were implemented with TensorFlow (v2.15) and Keras (v2.15) libraries. Code development was carried out using Jupyter Notebook (v6.5.4).

Experiments were conducted on a system equipped with an Intel Core i5-12400 CPU (6 cores, 12 threads) clocked at 2.50 GHz. Deep learning experiments were accelerated using Intel® UHD Graphics 730 memory. The system was equipped with 24 GB of DDR4 RAM. Data storage and retrieval were facilitated by a 500 GB NVMe SSD.

## Data Availability

The databases used can be asked on demand, they are ownership of the Centre for Biomedicine, Hull York Medical School, University of Hull.

